# Group interpersonal psychotherapy for depression in Uganda: intervention effects and the influence of social desirability bias in a pilot cluster randomized trial

**DOI:** 10.64898/2026.07.01.26357066

**Authors:** Elly Atuhumuza, Jacquellyn Nambi Ssanyu, Andrew Fraker, Shakirah Nakalungi, Morris Ndeezi, Vincent Mujune, Charlotte Oloya, Molly Naisanga, Jeffrey McManus, Crystal Huang, Roscoe Kasujja

**Author notes:** **Corresponding author:** Elly Atuhumuza, Address: StrongMinds, Kampala, Uganda.

## Abstract

**Introduction:** Group interpersonal psychotherapy (IPT-G) can expand access to depression care, but assessment conditions may influence outcomes. This pilot cluster-randomized trial was designed to refine outcome-measurement procedures and generate preliminary effect estimates to inform a larger trial. We evaluated a six-week model, examining associations between follow-up assessment conditions and depressive symptoms.

**Methods:** In Mayuge, Uganda, female participants aged ≥13 years with Patient Health Questionnaire-9 (PHQ-9) scores ≥10 were randomized by village to IPT-G or enhanced treatment as usual. Primary outcome was the PHQ-9 score. Assessments occurred at baseline, two weeks and three months post-treatment. During early two-week follow-up, therapy facilitators remained visible near interview locations while mobilizing participants; thereafter, they left before subsequent interviews. At three months, interviews were conducted independently and participants were randomly assigned to receive a prompt emphasizing confidentiality, accurate reporting and that incentives were not contingent on responses. We examined associations between these two conditions and PHQ-9 scores.

**Results:** Of 292 randomized participants, 263 provided three-month data. Mean PHQ-9 scores were lower in intervention than control participants at two weeks (1.73 versus 15.14; mean difference [MD] -13.41, 95% confidence interval [CI] -16.10 to -10.71) and three months (3.19 versus 10.97; MD -7.78, 95% CI -9.66 to -5.90; both p<0.001). Among intervention participants, prior assessment without facilitators and receipt of the confidentiality and honest-reporting prompt were jointly associated with 4.41-point higher PHQ-9 scores versus the reference condition (95% CI 2.37 to 6.45; p<0.001). In exploratory sensitivity analysis, an intervention subgroup exposed to both conditions had lower three-month PHQ-9 scores than controls (MD -5.19, 95% CI -7.39 to -2.99; p<0.001).

**Conclusions:** Six-week IPT-G was associated with significant symptom reductions, but follow-up procedures influenced the apparent benefit, highlighting the need for independent outcome assessment in community-delivered psychological intervention trials.

**Trial registration:** Pan African Clinical Trials Registry, PACTR202606549854263; retrospectively registered.

## Introduction

Depression is a leading cause of disability worldwide and a major contributor to years lived with disability, with the burden falling disproportionately on low- and middle-income countries (LMICs) [1], where an estimated 80% of people with mental disorders receive no treatment [2]. Uganda exemplifies this challenge: depression prevalence in the country is estimated to exceed 30% in some populations, driven by structural factors including poverty, food insecurity, gender-based violence, and high HIV/AIDS prevalence [3]. At the same time, the country has fewer than one psychiatrist per million people, who are also mainly concentrated in urban areas [4], with barriers such as stigma and low-income levels further limiting access to care [5,6]. Without radical investment in scalable, community-based approaches, this treatment gap will persist across generations [2].

Task-sharing, in which trained lay workers deliver defined components of mental health care with ongoing supervision and support from specialists, has emerged as a viable, evidence-supported strategy for narrowing this gap in LMICs [2]. Within this approach, brief structured psychological interventions that can be manualized, supervised and delivered by non-specialists are especially relevant. The World Health Organization’s Mental Health Gap Action Program identifies Interpersonal Psychotherapy (IPT) as a primary evidence-based treatment for depression appropriate for delivery within task-sharing frameworks [7].

Group Interpersonal Therapy (IPT-G), the group-adapted format, is particularly well-suited to resource-constrained settings because it extends reach, reduces per-patient cost and leverages community and relational dynamics that are central to mental health in many sub-Saharan African cultural contexts [8,9]. Foundational work in Uganda demonstrated that lay worker-delivered IPT-G could significantly reduce depressive symptoms and improve functioning relative to control conditions [8]. Verdeli et al. [9] simultaneously documented the adaptation process, showing that IPT’s interpersonal focus translated effectively into the Ugandan cultural context through deliberate modification of language, metaphors and problem area framing. These foundational studies prompted the broader uptake of IPT-G across sub-Saharan Africa.

StrongMinds, an organization dedicated to scaling mental health treatment, subsequently developed a further-adapted model that shortened the intervention from 16 weeks to six weeks to increase feasibility and reach. This model is delivered in weekly 90-minute group sessions by community lay workers, referred to as Therapy Facilitators (TFs), using a structured, manualized protocol. Recent evidence supports the promise of shorter and community-delivered IPT-G models. Program data from over 45,000 adults receiving this model in Uganda and Zambia show significant pre-to-post reductions in depressive symptoms [10], and a recent trial in central Uganda found a six-session IPT-G model with problem area concordant participants comparable in outcomes to an eight-session model with problem area discordant participants [11]. However, the six-week model has not been evaluated in a randomized trial in which groups were formed without deliberate matching by primary interpersonal problem area. Existing evidence relies heavily on routine programmatic data collected under conditions that may be vulnerable to systematic bias in self-reporting.

Trials of depression interventions typically rely on participant-reported symptom measures such as the Patient Health Questionnaire-9 (PHQ-9) [12]. In community-based implementation settings, responses to such measures may be shaped by features of the assessment context, including whether interviewers are perceived as affiliated with the intervention, whether confidentiality is emphasized, or whether facilitators are physically present during assessment [13,14]. These influences are variously described as social desirability bias, a form of response bias [15]. In trials utilizing self-reported measures like the PHQ-9, this bias typically manifests as a systematic under-reporting of depressive symptoms or an inflation of recovery rates post-intervention [16]. In some cultural contexts, these distortions can be amplified by the desire to avoid the stigma associated with persistent mental distress [17]. Such risks may be especially pronounced in LMIC settings, where the same proximity, trust, and relational dynamics that support community-based delivery may also blur the boundaries between care provision and evaluation.

Although concerns about social desirability bias in self-reported mental health measures are well documented, including in studies comparing interviewer-administered and self-administered modes of mental health assessment [13], examining whether respondents underreport sensitive PHQ-9 symptoms [18], and using cognitive interviewing to explore how respondents interpret and disclose symptoms [19], less is known about how assessment conditions shape estimated treatment effects in community-based mental health intervention trials.

This study was designed as a pilot cluster randomized trial to refine trial procedures, test outcome measurement protocols, and generate preliminary effect estimates to inform a larger randomized trial. During pilot implementation, variation in follow-up assessment conditions created an opportunity to examine whether reported PHQ-9 outcomes were sensitive to procedures expected to influence social desirability bias. The analysis therefore estimated the preliminary effect of the StrongMinds-adapted six-week IPT-G model on depressive symptoms and assessed whether reported symptoms differed according to two assessment conditions: whether therapy facilitators were present during assessment, and whether participants received an honesty and confidentiality prompt before outcome measurement. These analyses were intended to generate bias-informed estimates to guide interpretation of the pilot findings and strengthen the design of the forthcoming larger RCT.

## Materials and methods

### Setting

The study was done in Mayuge District, located in the Eastern Region of Uganda approximately 120 kilometres east of Kampala (the capital city) along the Uganda - Kenya highway. The district lies on the northern shores of Lake Victoria, with an estimated population of 570,000 people, predominantly rural [20]. The district falls within the Busoga subregion, one of Uganda’s high-poverty areas, with poverty driven by low household incomes, limited educational attainment and constrained access to health services. Formal mental health services in Mayuge, as in most Ugandan districts, are limited. District health planning in this region has historically depended on non-government partners to support mental health service delivery, with little integration of mental health into primary care. Study data were collected from June to November 2025.

### Study design

A pilot cluster randomized controlled trial design was used to evaluate the effects of a community-based six-week IPT-G intervention on depressive symptoms and to assess whether follow-up assessment conditions influenced reported outcomes.

### Intervention and comparator

Participants were randomly allocated to one of two study arms: an intervention arm, which received the six-week IPT-G program, or the Enhanced Treatment as Usual (ETAU) control arm, which included a one-time psychoeducation session on depression, PHQ-9 screening, and referral to local mental health services where appropriate.

The intervention arm received the six-week StrongMinds IPT-G program. The therapy is rooted in the IPT model, focusing on four primary problem areas that may be linked to the onset or maintenance of depression: grief, role disputes, role transitions and interpersonal deficits. The intervention was delivered in weekly group sessions designed to last approximately 90 minutes by TFs recruited from the local community. Under this program, TFs undergo an initial five-day training followed by weekly review sessions. The training covers the principles of IPT-G, group facilitation skills and mental health first aid. Groups typically comprise 8 to 12 eligible adolescents and women from the same village. The six-week model follows a structured, phased format, beginning with pre-group preparation and moving through an initial phase, middle phase and termination phase (Table 1). The standardized StrongMinds adaptation incorporates local idioms, examples and relational norms to support cultural relevance and acceptability in the Ugandan context.

**Table 1:**
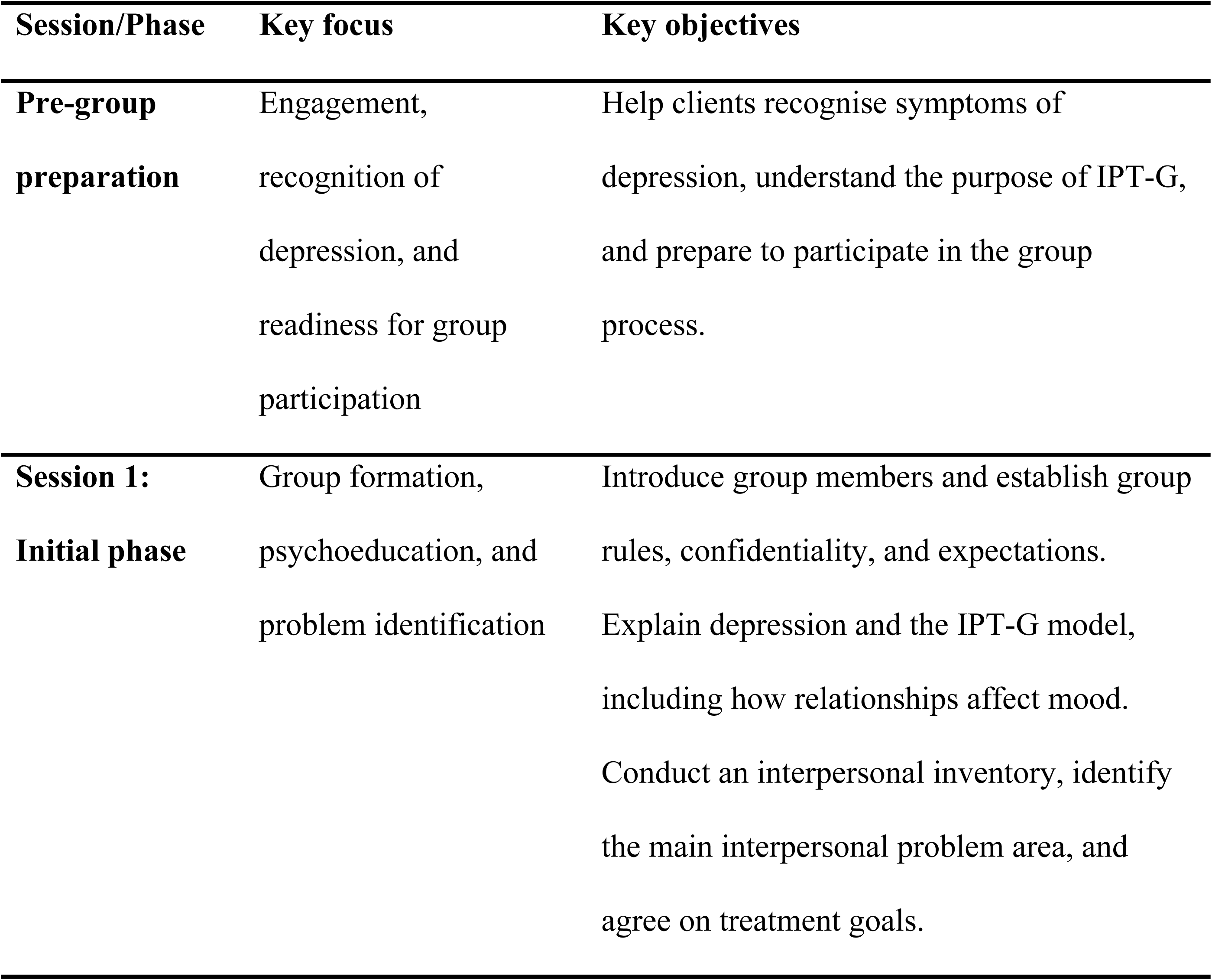
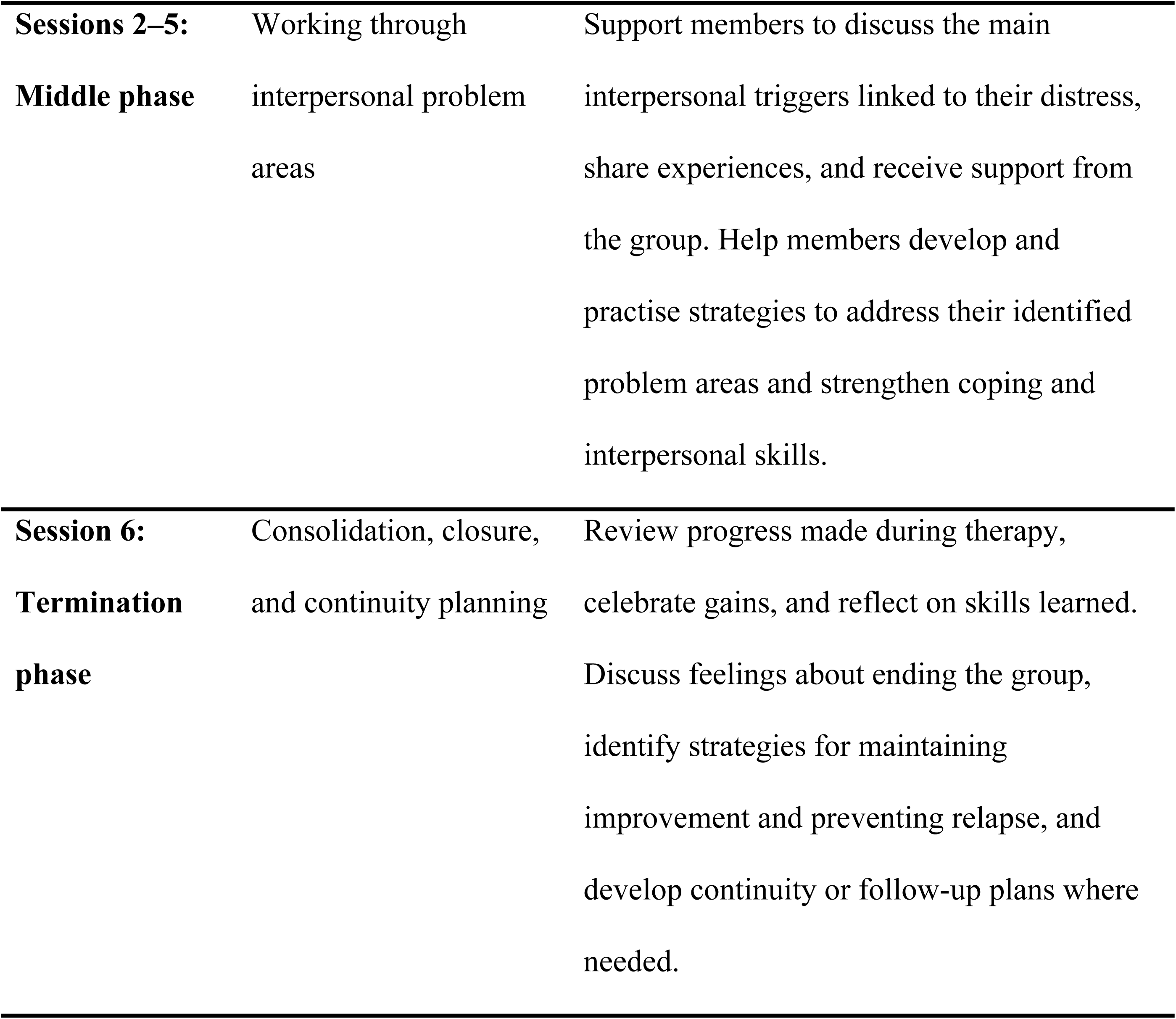
The IPT-G session structure.

### Participants

Participants were recruited through community mobilization conducted by StrongMinds facilitators, who sensitized residents about depression and invited interested individuals for screening. Initial pre-screening was conducted using the PHQ-4, a four-item instrument assessing depression and anxiety symptoms [21]. The PHQ-4 yields a total score ranging from 0 to 12, with scores commonly classified as none/minimal psychological distress (0 - 2), mild distress (3 - 5), moderate distress (6 - 8), and severe distress (9 - 12); higher scores indicate greater symptom burden. In line with StrongMinds’ programmatic screening procedures, individuals scoring 5 points or greater were invited to centralized locations, including schools and places of worship, for a formal research screening session held one to two weeks later.

At this session, trained research assistants administered the PHQ-9, a nine-item measure of depressive symptom severity with total scores ranging from 0 to 27, where higher scores indicate more severe depressive symptoms. Individuals scoring ≥10 on the PHQ-9, consistent with moderate to severe depressive symptoms, were considered eligible for the trial. Other eligibility criteria included being a permanent residence in the study area, female sex and being 13 years or older. Exclusion criteria included inability to provide consent or assent, withdrawal of consent or assent, suicidal ideation, or identification as a survivor of gender-based violence. Participants reporting suicidal ideation or gender-based violence were immediately referred for further support according to the study referral protocol.

### Sample size

A total of 24 villages were included in the trial, with 12 randomized to the intervention arm and 12 to ETAU. The initial target was to enrol 10 eligible participants per village, allowing a maximum of 12 participants per village. Each village formed one study cluster and contributed one group; villages were not combined to form groups. Half of the groups comprised young women and adolescents aged 24 years and younger (6 intervention, 6 ETAU) and half comprised adults older than 24 years (6 intervention, 6 ETAU). In total, 292 participants were randomized. As a pilot trial, the sample size was intended primarily to assess feasibility, refine study procedures, test outcome measurement and generate preliminary estimates to inform the design of a larger randomized trial.

### Randomization

Selected villages were randomly assigned to either the intervention or ETAU arm of the study using computer-generated random numbers. The village served as the unit of randomization, ensuring that all eligible individuals within each cluster received the same treatment status. This village-level randomization approach was used to minimize contamination between study arms and to align with the natural delivery structure of the intervention. Allocation of study arms was conducted independently from the field research team and concealed until after the baseline was completed.

### Blinding

Blinding of participants and intervention providers was not feasible due to the nature of the intervention. Outcome data were collected by trained research assistants who were not involved in intervention delivery. However, complete blinding of outcome assessors could not be guaranteed, particularly at follow-up, where participants may have disclosed their participation in the intervention.

### Outcomes

The primary outcome was depressive symptom severity, measured using the PHQ-9 at baseline, two weeks post-treatment and three months post-treatment. Intervention effects were estimated as differences in mean PHQ-9 scores between the intervention and control arms at follow-up. Three binary indicators were derived to aid clinical interpretation: remission, defined as a PHQ-9 score <5; treatment response, defined as a reduction of at least 50% from baseline; and minimal clinically important difference (MCID), defined as a reduction greater than 4 PHQ-9 points. A PHQ-9 score <5 corresponds to the none/minimal symptom range, while a 50% reduction and a five-point decline are commonly used indicators of clinically important improvement in depression treatment monitoring [12,22,23].

### Data collection procedures

Data were collected at three time points: baseline before treatment, two weeks post-treatment and three months post-treatment. All assessments were conducted by trained research assistants using structured, interviewer-administered questionnaires on electronic data collection tools.

#### Baseline/pre-treatment assessment

At baseline, research assistants collected socio-demographic and household characteristics, as well as pre-intervention depressive symptom severity using the PHQ-9, and other secondary outcomes not reported in this paper. Baseline data were collected after eligibility confirmation and consent or assent for study participation.

#### Two-week post-treatment assessment

The first follow-up assessment was conducted two weeks after completion of the six-week IPT-G intervention. Research assistants repeated the PHQ-9 and other outcome measures to assess early post-treatment changes in depressive symptoms.

During the initial five days of the two-week follow-up data collection, TFs supported participant mobilization by contacting participants and directing them to a central location within each village. In 176 of the 258 interviews conducted at this round, TFs remained visible, although out of earshot, while interviews were conducted. This facilitator involvement in mobilization was adopted for pragmatic reasons, as it reduced the time and cost of locating participants during the pilot. However, TF presence near the interview location was an unintentional departure from the intended independent assessment protocol and was identified only after data collection was underway.

Once identified, the procedure was revised. TFs could continue to support mobilization by contacting and directing participants to the interview location, but they were required to leave before interviews began. The remaining two-week follow-up interviews were therefore conducted without TFs present. This mid-round protocol variation was later treated analytically as a difference in assessment conditions, comparing interviews conducted with and without TF presence.

#### Three-month post-treatment assessment

The second follow-up assessment was conducted three months after completion of the intervention. Research assistants again administered the PHQ-9 and other outcome measures. By this stage, the revised independent assessment procedure was in place, and TFs were not present during interviews.

To reinforce confidentiality and reduce potential reporting pressure at the point of outcome measurement, a standardized prompt was introduced immediately before the outcome measurement sections at the three-month follow-up. The prompt emphasized that the research team was independent of TFs and StrongMinds program staff, that individual responses were confidential, that honest reporting was valued even if symptoms had worsened, that symptom trajectories after treatment could vary, and that participation incentives were guaranteed regardless of responses. The prompt was randomly assigned at the individual level using the built-in randomization function in KoboToolbox, generating approximately equal allocation to prompted and unprompted assessment conditions.

Together, the variation in TF presence at the two-week assessment and random assignment of the honesty prompt at three months allowed participants to be classified according to two assessment conditions: whether they had previously been assessed with TFs present, and whether they received the honesty prompt before three-month outcome measurement.

#### Research assistant training and participant safety

Research assistants received standardized training on questionnaire administration, ethical conduct, confidentiality and identification of participant distress. Participants who exhibited severe psychological distress or suicidal ideation during data collection were referred to appropriate mental health services according to a predefined referral protocol.

### Intervention fidelity monitoring

Fidelity to the StrongMinds six-week IPT-G model was supported through regular supervision and monitoring of TFs. Supervisory staff conducted scheduled observations of IPT-G sessions using standardized observation checklists that assessed adherence to the core IPT-G protocol, delivery of session content, time management, group facilitation skills and maintenance of a supportive and non-judgmental group environment. These procedures were intended to support fidelity while preserving the conditions of routine program delivery.

### Patient and public involvement

Patients and members of the public were not formally involved in the design of the study, selection of outcome measures, data analysis, interpretation of results, or preparation of this manuscript. The intervention was delivered through community-based structures, and local Therapy Facilitators supported community sensitisation and participant mobilisation. Study procedures were implemented in community locations selected for accessibility and privacy.

Participants were not involved in determining whether or how study findings would be disseminated. Aggregate findings will be shared with relevant community, district, and program stakeholders in a form that does not identify individual participants.

### Data analysis

Primary analyses followed the intention-to-treat principle, with participants analyzed according to the study arm to which their village was randomized, irrespective of intervention attendance or protocol adherence. Baseline balance analyses included all eligible randomized female participants. Main outcome analyses were conducted among participants with available three-month PHQ-9 data, which defined the primary analytic panel.

Baseline characteristics were summarized by study arm using means and standard deviations (SD) for approximately normally distributed continuous variables, medians and interquartile ranges (IQRs) for skewed continuous variables, and percentages for categorical variables. Balance between arms was assessed using standardized differences and regression-based p-values, with standard errors clustered at village level.

Primary treatment effects were estimated by comparing IPT-G and ETAU participants at each follow-up time point. For continuous outcomes, we estimated mean differences between study arms using linear regression. For binary outcomes, we estimated absolute percentage-point differences using linear probability models. All main treatment-effect models used village-level cluster-robust standard errors to account for the cluster-randomized design. The main full-sample outcome tables report unadjusted treatment contrasts at each follow-up point. This approach was used to keep the pilot results directly interpretable, particularly because baseline PHQ-9 and other key baseline characteristics were well balanced between study arms.

The social desirability bias protocol analyses examined whether reported PHQ-9 scores varied according to the two follow-up assessment conditions: absence of therapy facilitators during assessment and receipt of the honesty and confidentiality prompt. Because facilitator absence resulted from a mid-round protocol change rather than random allocation, we first assessed baseline comparability between participants assessed with and without facilitators present. We then estimated the association between each protocol condition and reported PHQ-9 scores, stratifying analyses by treatment arm and age group where relevant. These models adjusted for baseline PHQ-9 score and used village-level cluster-robust standard errors.

To examine the joint effect of the two protocol conditions, we estimated factorial models including treatment status, facilitator independence, the honesty and confidentiality prompt, and their interactions. These models included baseline PHQ-9 as a covariate. Finally, to estimate a more conservative primary treatment contrast under bias-minimizing assessment conditions, we compared the full control arm with the subgroup of intervention participants assessed both independently of facilitators and after receiving the honesty prompt. This comparison was adjusted for baseline PHQ-9 score using analysis of covariance.

To assess robustness of the primary three-month PHQ-9 estimate to loss to follow-up and analytic approach, we conducted baseline-adjusted complete-case analyses, inverse-probability-of-retention weighted analyses, village-level mean analyses, and wild-cluster bootstrap-t inference. Stabilized retention weights were estimated using treatment arm, baseline PHQ-9 score, age, age group, and marital status, and were truncated at the 1^st^ and 99^th^ percentiles. Village-level analyses used mean PHQ-9 scores adjusted for mean baseline PHQ-9 and weighted by the number of participants contributing outcome data per village. Wild-cluster bootstrap-t inference used 9,999 replications. These analyses were treated as sensitivity checks rather than the primary basis for inference. All analyses were conducted in Stata 18, with p<0.05 interpreted as statistically significant.

### Ethical considerations

Informed consent was obtained from all eligible participants before enrolment. Given varying literacy levels, the consent process was conducted orally in each participant’s preferred local language, with research assistants reading the study objectives, procedures, risks, benefits, confidentiality protections, and participant rights aloud before written consent was documented. Adult participants provided written informed consent. For participants below 18 years, written parental or guardian consent and participant assent were obtained. Participants below 18 years were considered emancipated minors if they were married, pregnant, had a child, or were responsible for their own livelihood; in such cases, written informed consent was obtained directly from the participant. All participants were informed that participation was voluntary and that they could withdraw at any time without any effect on their access to current or future health care or other services.

Participants were screened to exclude those reporting suicidal ideation, identifying as survivors of gender-based violence, and both were immediately referred for further screening and treatment. For participants who exhibited signs of distress, a protocol for immediate referral to a qualified mental health professional or facility was implemented, with follow-up conducted within 48 to 72 hours. To protect participant privacy, all data were de-identified using unique alphanumeric codes. No personally identifiable information was included in the study datasets. All digital records were stored on password-protected, encrypted servers accessible only to the principal investigator and the core research team. In recognition of their time, participants were provided with a modest compensation package consisting of sugar and soap. The nature of this compensation was approved by the local ethics committees as being culturally appropriate and non-coercive, aligned with standard research practices in the region. Ethical approval for this study was granted by the Mildmay Research and Ethics Committee (Reference Number MUREC-2025-799) and Uganda National Council of Science and Technology (Reference Number SS3945ES).

### Trial registration

This pilot cluster-randomized trial was retrospectively registered in the Pan African Clinical Trials Registry (PACTR202606549854263) on 29 June 2026. Participant recruitment began in June 2025. The study was initiated as a program-embedded pilot intended to refine intervention delivery, recruitment procedures, outcome measurement, follow-up processes, and sample-size assumptions for a planned definitive trial. Although ethical approval was obtained before recruitment, trial registration was not completed before participant recruitment began. This was an administrative oversight. Nonetheless, the primary outcome and core study procedures were specified in the ethics-approved protocol (Supplementary File S2) before recruitment. Deviations and subsequent refinements to follow-up assessment procedures are reported transparently in this manuscript and to the research and ethics committee.

## Results

### Participant flow

A total of 418 individuals were assessed for eligibility, of whom 126 were excluded, primarily because they did not meet the inclusion criteria (n=116), while 10 were referred due to suicidal ideation (Fig 1). The remaining 292 participants were randomized at cluster level, with 148 allocated to the intervention arm and 144 to the ETAU control arm. In the intervention arm, 141 participants (95.3%) attended at least one IPT-G session, while 7 did not receive the allocated intervention, including 4 who never attended and 3 due to clerical errors. By the three-month follow-up, 17 participants in the intervention arm and 12 in the control arm had been lost to follow-up. In addition, 7 intervention participants discontinued treatment, defined as attending fewer than four sessions. Participants who did not receive or discontinued the intervention were not excluded from follow-up or analysis on that basis; they remained in their originally randomized arm and were included in the intention-to-treat analysis where follow-up outcome data were available. Three-month PHQ-9 data were available for 131 of 148 intervention participants (88.5%) and 132 of 144 control participants (91.7%). Retention was 3.2 percentage points lower in the intervention arm, but there was no evidence of differential retention by study arm (95% CI: -11.3 to 5.0; p=0.431).

**Fig 1:**
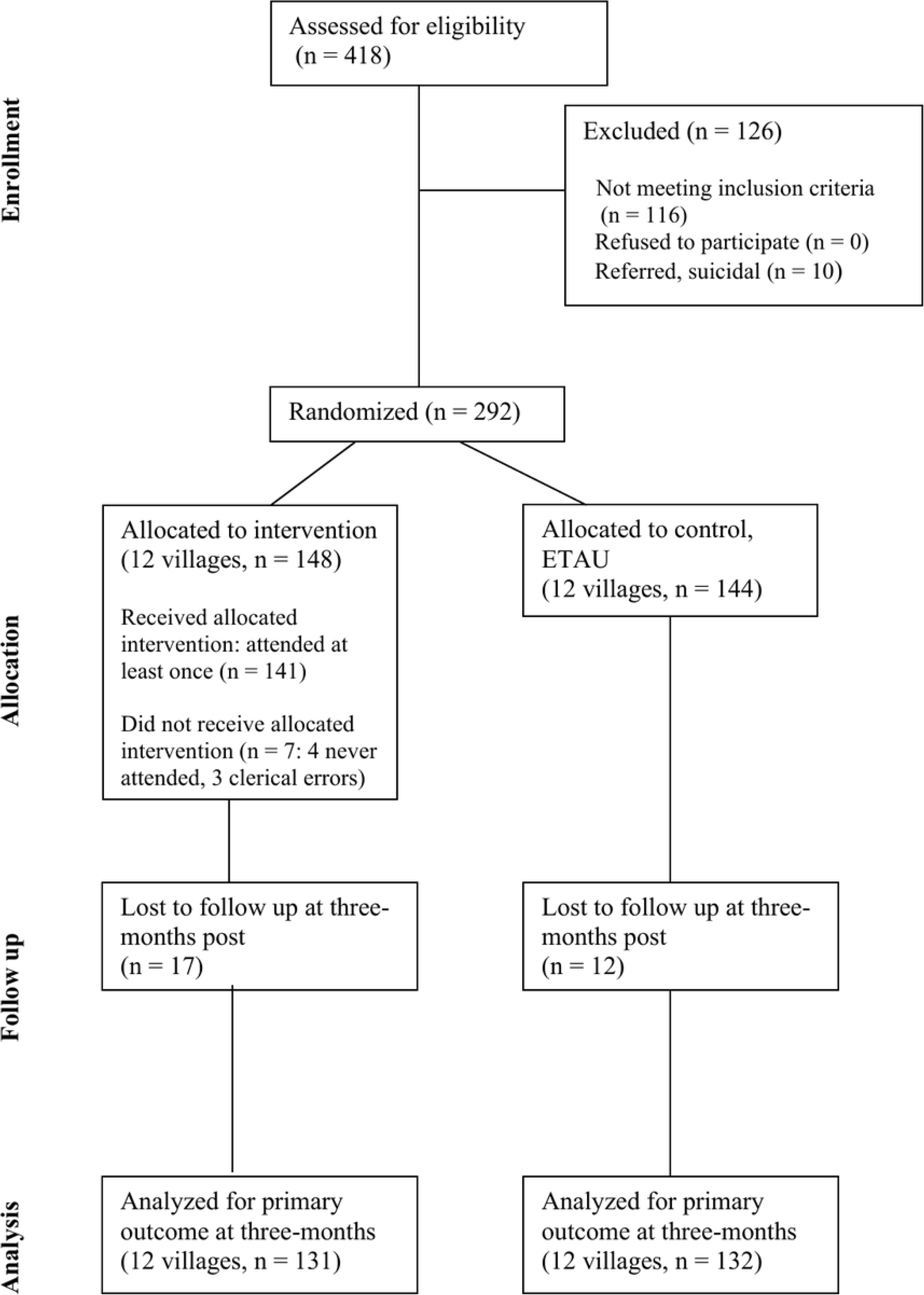
Participant Flow into the study.

### Intervention delivery and fidelity

Participant exposure to the intervention was high. Among participants randomized to the intervention arm, average attendance was 5 of 6 sessions; 95.3% attended at least one session, 93.1% attended at least three sessions, 81.9% attended at least five sessions and 49.3% completed all six sessions.

### Baseline characteristics

Baseline characteristics were broadly comparable between study arms in both the full baseline sample and the three-month analytic panel (Table 2). In the full baseline sample, the median age was 27.5 years (IQR: 22.0 - 43.5; range: 16-80) in the intervention arm and 31.5 years (IQR: 22.0-50.0; range: 16-85) in the control arm. The proportion of adolescents and young women aged 16-24 years was also similar across arms: 47.3% in the intervention arm and 43.1% in the control arm. The two groups were comparable with respect to marital status, educational attainment, number of children, and baseline depressive symptom severity. Mean baseline PHQ-9 scores were nearly identical in the two groups, at 15.62 in the intervention arm and 15.60 in the control arm (p=0.983). This baseline comparability was retained in the three-month analytic panel.

**Table 2:** Baseline characteristics of all randomized participants and the three-month.

| Variable | Full baseline sample |  |  | Three-month panel sample |  |  |
| --- | --- | --- | --- | --- | --- | --- |
|  | Treatment<br>(N=148) | Control<br>(N=144) | p-value | Treatment<br>(N=131) | Control (N=132) | p-value |
| Age, median [IQR],<br>range | 27.5 [22.0-43.5],<br>16-80 | 31.5 [22.0-50.0],<br>16-85 | 0.541 | 29.0 [23.0-47.0],<br>16-80 | 31.5 [22.0-50.0],<br>16-85 | 0.645 |
| Younger participants |  |  |  |  |  |  |
| (16-24 years) | 47.3% | 43.1% | 0.824 | 43.5% | 42.4% | 0.954 |
| Married | 41.9% | 50.0% | 0.308 | 43.5% | 52.3% | 0.273 |
| Upper primary |  |  |  |  |  |  |
| education | 48.0% | 44.4% | 0.605 | 51.1% | 46.2% | 0.505 |
| Number of children | 3.59 (2.80) | 3.77 (2.85) | 0.848 | 3.74 (2.86) | 3.83 (2.89) | 0.934 |
| PHQ-9 score (SD) | 15.62 (4.28) | 15.60 (3.92) | 0.983 | 15.71 (4.17) | 15.55 (3.82) | 0.835 |

### Overall treatment estimates before accounting for assessment-condition variation

In the pooled intention-to-treat analysis, the intervention was associated with large improvements in depressive symptoms before accounting for variation in assessment conditions. At the two-week post-treatment follow-up, mean PHQ-9 scores were 1.73 in the intervention arm compared with 15.14 in the control arm, corresponding to an unadjusted mean difference of -13.41 points (95% CI: -16.10 to -10.71; p<0.001) (Table 3). This was accompanied by substantially higher remission, MCID achievement, and response rates in the intervention arm. Remission was achieved by 81.0% of participants in the intervention arm compared with 2.4% in the control arm; 92.2% achieved an MCID compared with 23.6%; and 80.2% achieved response compared with 10.6%.

**Table 3:** Primary outcomes under the initial follow-up protocols.

| Variable | Treatment<br>(N=131) | Control<br>(N=132) | Effect (95% CI) | p-value |
| --- | --- | --- | --- | --- |
| PHQ-9 |  |  |  |  |
| Baseline | 15.71 (4.17) | 15.55 (3.82) | 0.16 (-1.31 to 1.62) | 0.835 |
| 2 weeks | 1.73 (3.17) | 15.14 (5.69) | -13.41 (-16.10 to -10.71) | <0.001 |
| 3 months | 3.19 (3.86) | 10.97 (5.32) | -7.78 (-9.66 to -5.90) | <0.001 |
| Score change (PHQ-9) |  |  |  |  |
| 2 weeks | 14.12 (5.79) | 0.54 (5.36) | 13.58 (10.77 to 16.40) | <0.001 |
| 3 months | 12.52 (5.67) | 4.58 (5.32) | 7.94 (5.53 to 10.34) | <0.001 |
| Remission (PHQ-9 < 5) |  |  |  |  |
| 2 weeks | 81.0% | 2.4% | 78.6% (63.2 to 94.0) | <0.001 |
| 3 months | 71.0% | 7.6% | 63.4% (50.6 to 76.3) | <0.001 |
| MCID (>4 pts PHQ-9 reduction) |  |  |  |  |
| 2 weeks | 92.2% | 23.6% | 68.7% (52.4 to 84.9) | <0.001 |
| 3 months | 92.4% | 55.3% | 37.1% (25.6 to 48.5) | <0.001 |
| Response rate (50% reduction) |  |  |  |  |
| 2 weeks | 80.2% | 10.6% | 69.5% (53.6 to 85.5) | <0.001 |
| 3 months | 85.5% | 28.8% | 56.7% (42.6 to 70.9) | <0.001 |

By three months, differences between arms had attenuated but remained large. Mean PHQ-9 scores were 3.19 in the intervention arm and 10.97 in the control arm, corresponding to an unadjusted mean difference of -7.78 points (95% CI: -9.66 to -5.90; p<0.001). Remission remained higher in the intervention arm than in the control arm (71.0% versus 7.6%), as did MCID achievement (92.4% versus 55.3%) and response rates (85.5% versus 28.8%).

The three-month treatment estimate was robust to alternative analytic approaches (S3 Table). The baseline-adjusted complete-case estimate was -7.82 PHQ-9 points (95% CI: -9.84 to - 5.79), compared with -7.75 points in the inverse-probability-weighted analysis (95% CI: -9.78 to -5.73) and -7.77 points in the village-level analysis (95% CI: -9.86 to -5.68). Wild-cluster bootstrap-t inference yielded a 95% confidence interval of -9.88 to -5.70. All analyses remained statistically significant at p<0.001.

Age-stratified analyses showed lower mean PHQ-9 scores in the intervention than control arm at both follow-up time points among adults aged ≥25 years and younger participants aged 16–24 years (Fig 2). Among adults, mean PHQ-9 scores at two weeks were 0.60 in the intervention arm and 16.56 in the control arm, corresponding to an unadjusted mean difference of −15.96 points (95% CI: -18.43 to −13.50; p<0.001). At three months, mean scores were 2.49 and 11.61, respectively, corresponding to a difference of -9.12 points (95% CI: -11.24 to -6.99; p<0.001). Among younger participants, mean PHQ-9 scores at two weeks were 3.18 in the intervention arm and 13.06 in the control arm, corresponding to a difference of -9.88 points (95% CI: -13.70 to -6.07; p<0.001). At three months, mean scores were 4.11 and 10.11, respectively, corresponding to a difference of -6.00 points (95% CI: -8.72 to -3.28; p<0.001).

**Fig 2:**
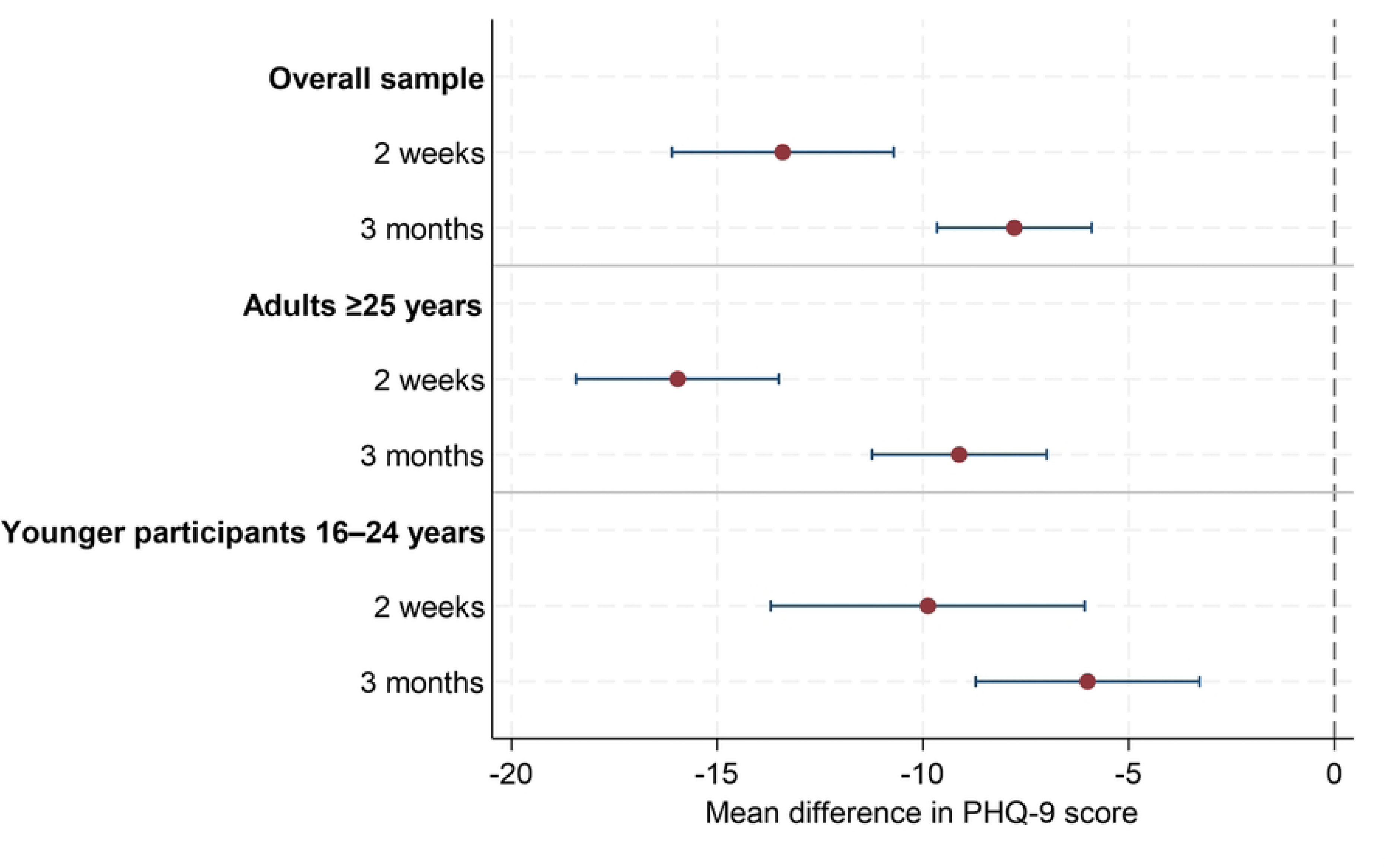
Unadjusted differences in mean PHQ-9 scores between intervention and control arms, overall and by age group.

In exploratory baseline-adjusted interaction models, the intervention effect at two weeks was larger among adults (-15.99 points; 95% CI: -18.41 to -13.57) than younger participants (-9.80 points; 95% CI: -13.82 to -5.78), with a 6.18-point difference in treatment effects (95% CI: 1.84 to 10.53; interaction p=0.007). At three months, the adjusted intervention-control difference was -9.10 points among adults (95% CI: -11.47 to -6.72) and -6.13 points among younger participants (95% CI: -8.93 to -3.33). The difference in treatment effects was smaller and did not meet conventional statistical significance at three months (2.97 points; 95% CI: - 0.51 to 6.45; interaction p=0.091).

### Effects of follow-up assessment conditions on PHQ-9 outcomes

#### Distribution of participants across protocol conditions

Among participants with three-month PHQ-9 outcome data, 165 had been assessed with TFs present or visible near the assessment location during the two-week follow-up, while 98 had been assessed without TFs present. Baseline characteristics were broadly similar across the two groups, although some imbalances were observed (Table 4). Participants assessed without TFs present were younger, with a median age of 24 years (IQR: 21-40; range: 16-80), compared with 35 years (IQR: 23-50; range: 16-85) among those assessed with TFs present. A higher proportion of the TF-absent group were younger participants aged 16-24 years (57.1% versus 34.5%; standardized difference 0.47). The TF-absent group also had a lower mean baseline PHQ-9 score than the TF-present group (14.79 versus 16.13; standardized difference -0.34; p=0.036). Smaller differences were observed for treatment arm allocation, upper primary education, and number of children, while marital status was nearly identical across groups.

**Table 4:** Baseline characteristics for the facilitator independence protocol change.

|  | TFs absent | TFs present | Standardized |  |
| --- | --- | --- | --- | --- |
| Variable | (n=98) | (n=165) | Difference | p-value |
| Treatment arm | 54 (55.1%) | 77 (46.7%) | 0.17 | 0.673 |
|  | 24.0 (21.0-40.0); | 35.0 (23.0-50.0); |  |  |
| Age, median [IQR]; range | 16-80 | 16-85 | -0.38 | 0.230 |
| Younger participants, 16-24 years | 56.0 (57.1%) | 57 (34.5%) | 0.47 | 0.207 |
| Married | 47.0 (48.0%) | 79 (47.9%) | 0.00 | 0.990 |
| Upper primary education | 50.0 (51.0%) | 78 (47.3%) | 0.08 | 0.512 |
| Number of children | 3.25 (2.92), n=76 | 4.11 (2.80), n=124 | -0.30 | 0.400 |
| Baseline PHQ-9 score | 14.79 (3.91) | 16.13 (3.97) | -0.34 | **0.036** |

**Table 5:** Distribution of participants across assessment conditions among participants with three-month PHQ-9 data.

| TF absent at two-week<br>assessment | Not prompted at three<br>months | Prompted at three<br>months | Total |
| --- | --- | --- | --- |
| No | 79 (30.0%) | 86 (32.7%) | 165 (62.7%) |
| Yes | 47 (17.9%) | 51 (19.4%) | 98 (37.3%) |
| Total | 126 (47.9%) | 137 (52.1%) | 263 (100.0%) |

At the three-month follow-up, the honesty and confidentiality prompt was randomized at the individual level, with approximately equal allocation: 126 participants (48%) did not receive the prompt, while 137 (52%) received it. Cross-classifying the earlier two-week TF-presence condition with three-month prompt assignment, 79 participants had been assessed with TFs present at two weeks and did not receive the prompt at three months, 86 had been assessed with TFs present and received the prompt, 47 had been assessed without TFs present and did not receive the prompt, and 51 had been assessed without TFs present and received the prompt.

#### Association between two-week facilitator presence and reported PHQ-9 outcomes

Assessment condition at the two-week follow-up showed different associations with reported PHQ-9 scores in the control and intervention arms. Among participants with three-month PHQ-9 data and non-missing two-week PHQ-9 scores, those assessed without TFs present had lower mean PHQ-9 scores overall than those assessed with TFs present, although the overall difference was not statistically significant (7.04 against 9.35; adjusted difference -2.15 points, 95% CI: -7.45 to 3.15; p=0.410) (S4 Table).

In the control arm, participants assessed without TFs present reported lower PHQ-9 scores than those assessed with TFs present (10.57 versus 16.95; adjusted difference -5.76 points, 95% CI: -8.78 to -2.73; p=0.002). In contrast, intervention participants assessed without TFs present reported higher PHQ-9 scores than those assessed with TFs present, although the confidence interval included the null (3.87 versus 0.65; adjusted difference 3.06 points, 95% CI: -0.09 to 6.21; p=0.056). This pattern was most pronounced among younger intervention participants aged 16–24 years, where assessment without TFs present was associated with a 5.08-point higher PHQ-9 score (6.50 versus 1.36; 95% CI: 3.00 to 7.16; p=0.001).

By the three-month follow-up, all interviews were conducted without TFs present. The three-month analysis therefore examined whether the earlier two-week assessment condition was associated with later PHQ-9 reporting. Earlier assessment without TFs present was no longer associated with reported PHQ-9 scores in the control arm overall (10.25 versus 11.33; adjusted difference -0.40 points, 95% CI: -3.13 to 2.32; p=0.750) (S5 Table). In the intervention arm, however, participants who had been assessed without TFs present at two weeks reported higher three-month PHQ-9 scores than those previously assessed with TFs present (4.69 versus 2.14; adjusted difference 2.62 points, 95% CI: 0.27 to 4.97; p=0.032). This pattern was observed in both age subgroups, although the estimate was statistically significant only among adults older than 24 years.

#### Effect of the honesty and confidentiality prompt

The honesty and confidentiality prompt showed a similar pattern to facilitator independence. At three months, the prompt was not meaningfully associated with PHQ-9 scores in the control arm, where prompted and unprompted participants reported similar symptom levels (S6 Table). The adjusted mean difference in the control arm was 0.15 points (95% CI: -1.31 to 1.61; p=0.825). In contrast, prompted participants in the intervention arm reported higher PHQ-9 scores than unprompted participants (4.07 versus 2.12; adjusted difference 1.95 points, 95% CI: 0.90 to 3.00; p=0.002). This pattern was observed among both younger intervention participants and adults aged ≥25 years. Among younger participants, prompted participants reported PHQ-9 scores 2.40 points higher than unprompted participants (5.23 versus 2.77; 95% CI: 0.20 to 4.59; p=0.036). Among adults, prompted participants reported PHQ-9 scores 1.56 points higher than unprompted participants (3.20 versus 1.61; 95% CI: 0.40 to 2.72; p=0.016).

#### Joint analysis of assessment conditions

Within each study arm, the reference group comprised participants assessed with TFs present at the two-week follow-up who did not receive the three-month honesty and confidentiality prompt. Among control participants, observed mean three-month PHQ-9 scores were broadly similar across assessment-condition combinations, ranging from 9.46 to 11.68 (Fig 3). Relative to the reference mean of 11.68, adjusted differences were -1.83 points among participants previously assessed without TFs present and not prompted (p=0.166), -0.67 points among prompted participants previously assessed with TFs present (p=0.423), and -0.01 points among those exposed to both conditions (p=0.993).

**Fig 3.**
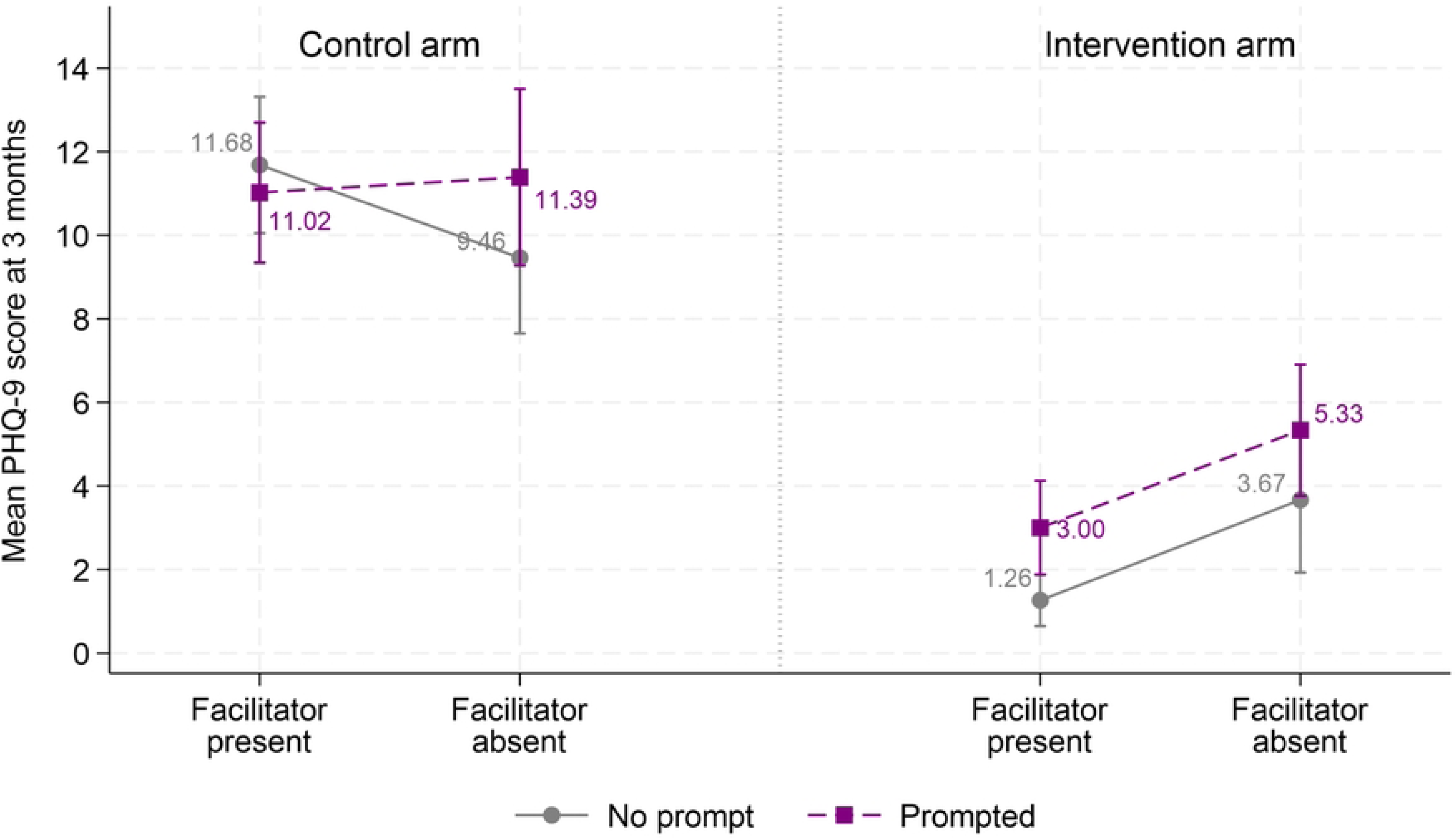
Observed mean three-month PHQ-9 scores by two-week facilitator-presence condition and three-month honesty and confidentiality prompt assignment, stratified by study arm. Points represent observed mean PHQ-9 scores and error bars represent 95% confidence intervals. The reference condition comprised participants assessed with Therapy Facilitators present at the two-week follow-up who did not receive the three-month honesty and confidentiality prompt.

Among intervention participants, observed mean PHQ-9 scores increased from 1.26 in the reference group to 3.67 among those previously assessed without TFs present and not prompted, 3.00 among prompted participants previously assessed with TFs present, and 5.33 among participants exposed to both conditions. Relative to the reference group, adjusted differences were 2.53 points (p=0.053), 1.56 points (p=0.016), and 4.41 points (p<0.001), respectively (Fig 3; S7 Table).

#### Treatment estimates under bias-minimizing assessment conditions

Under assessment conditions expected to reduce reporting pressure, the estimated treatment effect at three months was attenuated but remained substantial. In the full analytic sample, the unadjusted mean difference in PHQ-9 score between intervention and control participants at three months was -7.78 points. In a sensitivity analysis restricting the intervention group to participants who had been assessed without TFs present at the two-week follow-up and who received the honesty and confidentiality prompt at three months, the adjusted mean difference compared with the full control arm was -5.19 points (95% CI: -7.39 to -2.99; p<0.001) (Table 6).

**Table 6:**
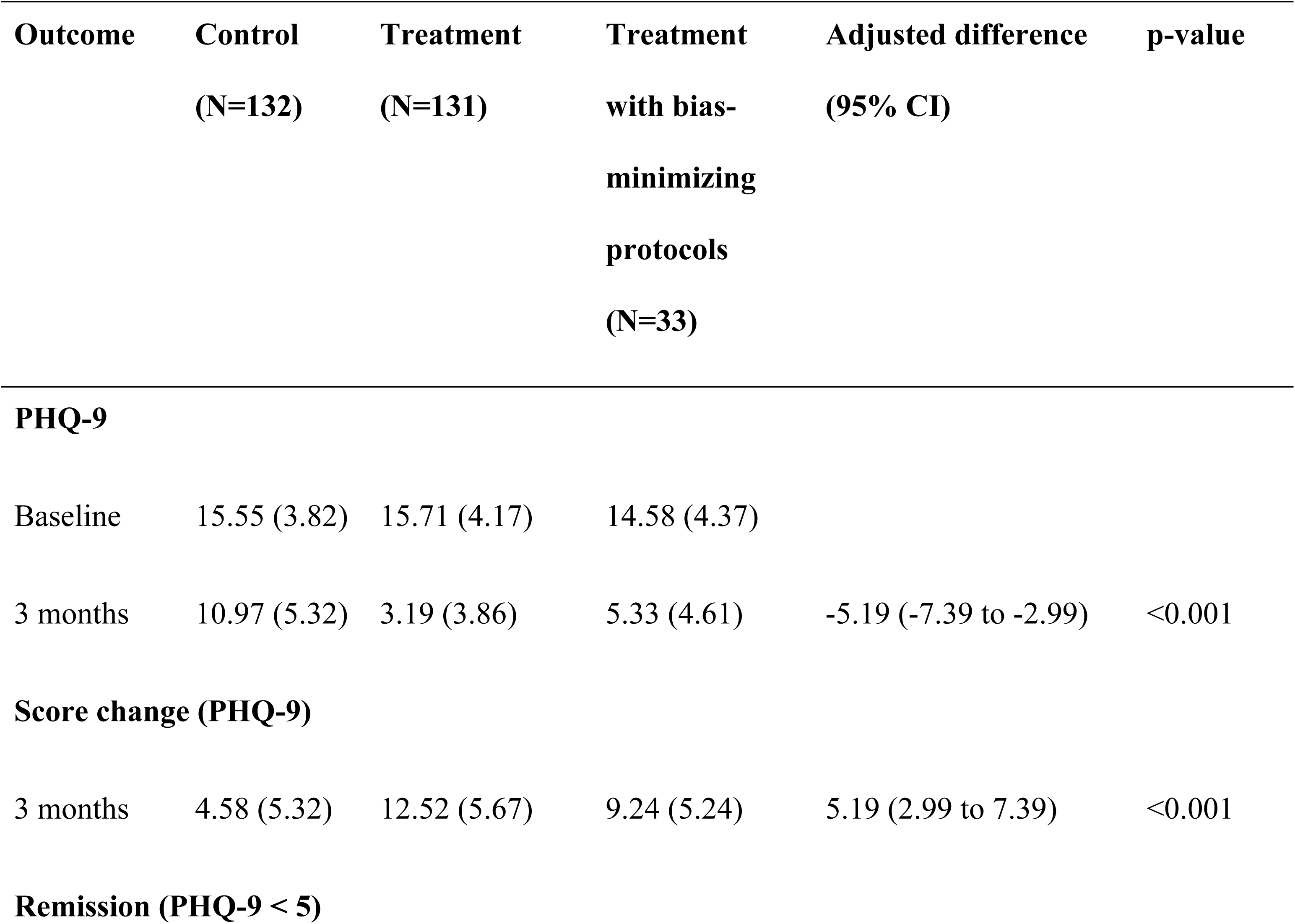
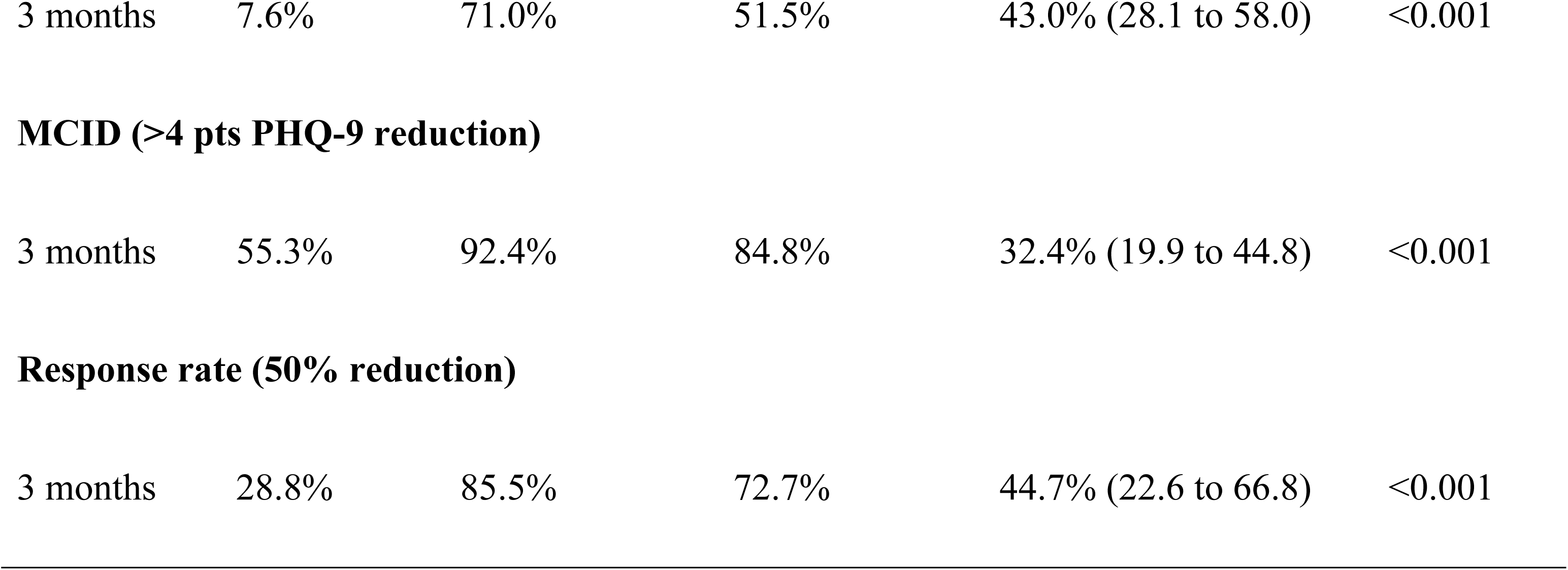
Three-month treatment estimates using the intervention subgroup assessed under bias-informed conditions.

| Outcome | Control<br>(N=132) | Treatment<br>(N=131) | Treatment<br>with bias-minimizing<br>protocols<br>(N=33) | Adjusted difference<br>(95% CI) | p-value |
| --- | --- | --- | --- | --- | --- |
| **PHQ-9** |  |  |  |  |  |
| Baseline | 15.55 (3.82) | 15.71 (4.17) | 14.58 (4.37) |  |  |
| 3 months | 10.97 (5.32) | 3.19 (3.86) | 5.33 (4.61) | -5.19 (-7.39 to -2.99) | <0.001 |
| **Score change (PHQ-9)** |  |  |  |  |  |
| 3 months | 4.58 (5.32) | 12.52 (5.67) | 9.24 (5.24) | 5.19 (2.99 to 7.39) | <0.001 |
| **Remission (PHQ-9 < 5)** |  |  |  |  |  |
| 3 months | 7.6% | 71.0% | 51.5% | 43.0% (28.1 to 58.0) | <0.001 |
| **MCID (>4 pts PHQ-9 reduction)** |  |  |  |  |  |
| 3 months | 55.3% | 92.4% | 84.8% | 32.4% (19.9 to 44.8) | <0.001 |
| **Response rate (50% reduction)** |  |  |  |  |  |
| 3 months | 28.8% | 85.5% | 72.7% | 44.7% (22.6 to 66.8) | <0.001 |

Clinical response indicators showed a similar pattern. Among intervention participants in this bias-informed subgroup, 17 of 33 participants (51.5%) met remission criteria, compared with 10 of 132 control participants (7.6%), corresponding to an adjusted difference of 43.0 percentage points (95% CI: 28.1 to 58.0; p<0.001). MCID achievement was also higher in the bias-informed intervention subgroup than in the control arm: 28 of 33 participants (84.8%) versus 73 of 132 participants (55.3%), with an adjusted difference of 32.4 percentage points (95% CI: 19.9 to 44.8; p<0.001). Response was similarly higher: 24 of 33 participants (72.7%) in the bias-informed intervention subgroup versus 38 of 132 participants (28.8%) in the control arm, corresponding to an adjusted difference of 44.7 percentage points (95% CI: 22.6 to 66.8; p<0.001).

## Discussion

This study found that the adapted six-week IPT-G model was associated with significant reductions in depressive symptoms. However, reported outcomes varied systematically according to follow-up assessment conditions. Both facilitator presence during assessment and the absence of an honesty and confidentiality prompt were associated with lower reported PHQ-9 scores in the intervention arm, suggesting inflation of symptom improvement under standard protocols. Importantly, even under these more conservative assessment conditions, IPT-G remained associated with better PHQ-9 outcomes, including higher remission, MCID achievement, and response rates than the control arm. These findings provide preliminary evidence supporting the potential value of the six-week IPT-G model, while also showing that measurement context can shape reported outcomes in community-based psychological intervention trials.

Under the initial follow-up procedures, IPT-G was associated with large reductions in depressive symptoms at both two weeks and three months. Although the estimated three-month difference was smaller among intervention participants assessed under conditions expected to reduce reporting pressure, a substantial treatment difference remained. This pattern suggests that the observed improvements were not wholly explained by follow-up procedures. These results are consistent with evidence from Uganda and other sub-Saharan African settings showing that lay worker-delivered IPT-G can reduce depressive symptoms when adapted for community delivery [10,11,24], and with six-month follow-up findings from rural Uganda showing that benefits of IPT-G can persist for several months beyond treatment completion [25]. The exploratory adjusted three-month difference of 5.19 PHQ-9 points was numerically larger than the 1.0-point difference reported at three months in a Group Problem Management Plus trial in Nepal [26]. However, differences in participant populations, baseline symptom severity, intervention content and dose, comparator conditions, and outcome-assessment procedures preclude conclusions about comparative effectiveness. Qualitative evidence from rural Uganda suggests that IPT-G may support recovery through social comparison, thought modification and mutual group support [27]. The six-week model may therefore provide a structured setting in which participants can discuss interpersonal difficulties, receive support, and develop strategies for managing distress.

Exploratory analyses suggested that the early post-treatment effect was smaller among younger participants aged 16–24 years than among adults, although both groups showed substantial reductions in depressive symptoms. By three months, the difference between age groups was smaller and did not meet conventional statistical significance. These findings should be interpreted cautiously because the pilot was not powered for subgroup analyses and younger participants were more commonly assessed under the revised facilitator-independent protocol. Future, adequately powered trials should examine whether this pattern reflects differences in treatment response, intervention fit, or outcome reporting conditions.

The attenuation observed under bias-minimizing assessment conditions is, however, notable. The findings suggest that social desirability bias may have influenced reported outcomes, particularly in the intervention arm. Participants assessed in the presence, or perceived proximity, of therapy facilitators may have felt pressure to report improvement out of gratitude, loyalty, fear of disappointing facilitators, or concern that negative responses could affect future support. The honesty and confidentiality prompt appeared to reduce this pressure by separating truthful reporting from continued access to incentives or services. Similar reporting biases have been documented in measures of depression and other sensitive or stigmatized behaviours, including self-reported drug and alcohol use among people living with HIV [13,17,28,29].

The weaker pattern in the control arm suggests that these differences were not simply a general survey effect, but may have been related to participants’ experience of receiving the intervention. This has implications for community-delivered psychological interventions, where the relational features that support engagement and therapeutic change may also heighten expectations to report improvement. Overall, the findings indicate a meaningful treatment signal, but one whose estimated magnitude is sensitive to how, where and by whom outcomes are measured.

The finding that earlier two-week TF presence was associated with three-month PHQ-9 reporting should be interpreted cautiously. Unlike the two-week analysis, where TF presence and outcome assessment occurred at the same time, all three-month assessments were conducted without TFs present. The three-month association therefore cannot be interpreted as a direct same-round facilitator-presence effect. One possible explanation is assessment-history or panel-conditioning: earlier assessment experiences may shape later reporting by influencing participants’ trust in the research process, expectations about desirable responses, or consistency with prior answers. However, the evidence for persistence of social desirability effects after removal of the immediate trigger is limited, particularly in community-based mental health trials. Alternative explanations include residual confounding from the non-randomized timing of the two-week protocol change, differences in participant composition, or true differences in symptom trajectories. We therefore interpret this as an exploratory association rather than evidence of a persistent facilitator-presence effect.

The findings from this study have direct implications for the design and interpretation of community-delivered psychological intervention trials in LMIC settings, where task-shared models are both most needed and most susceptible to the conditions that can generate social desirability bias. The present study offers experimental quantification of this susceptibility. A 4.41-point upward shift in PHQ-9 scores among intervention participants assessed under both bias-minimizing conditions, with no equivalent pattern in the control arm, suggests that assessment context can influence reported outcomes in the direction of apparent benefit, even when standard consent and confidentiality procedures are in place. These dynamics, including facilitator proximity and unclear separation of program and research roles, are not unique to this study and may operate to varying degrees across community-based trials more broadly, making bias-reduction strategies a worthwhile investment. Independent outcome assessment should therefore be prioritised, particularly when outcomes are self-reported and intervention delivery depends on trusted community-based facilitators. Simple procedural safeguards may be especially valuable, such as ensuring facilitators are not present during assessments, clearly separating program staff from research staff, reinforcing confidentiality, and making it explicit that incentives or future support do not depend on reporting improvement. These are low-cost adjustments, but their effect on estimates may be significant.

The study has several strengths. It was embedded in a real-world community delivery model, making the findings directly relevant to implementation settings where task-shared psychological interventions are likely to be scaled. The study also captured variation in assessment conditions that are rarely measured directly, even though such conditions are common in pragmatic trials and routine program evaluations. The individual randomization of the honesty and confidentiality prompt strengthens the interpretation of that component, while the comparison of multiple assessment conditions provides a broader view of how reporting bias may operate. As a result, the study contributes not only preliminary evidence on the six-week IPT-G model, but also practical evidence on how outcome measurement procedures can shape estimated intervention effects.

Nonetheless, several limitations should be considered when interpreting these findings. This pilot trial was designed to refine trial procedures, test outcome measurement protocols, and generate preliminary estimates to inform the design of a forthcoming, fully powered cluster randomized trial; it was not intended to provide definitive estimates of intervention effectiveness, subgroup differences, or assessment-condition mechanisms. The study enrolled females only and was conducted in rural communities in Mayuge District, Uganda. The findings may therefore not be generalizable to men or to populations in other settings. Although the intervention was randomized at cluster level, facilitator presence during follow-up was not randomized and resulted from a mid-round protocol change. Participants assessed with and without facilitators present differed on some baseline characteristics, including adolescent composition and baseline PHQ-9 scores; residual confounding therefore cannot be ruled out. The subgroup assessed under both bias-minimizing conditions was also small (n=33), limiting precision and making subgroup estimates, particularly by age, exploratory.

The forthcoming larger RCT will prospectively incorporate independent outcome assessment and pre-specified assessment-condition procedures from the outset. It will be adequately powered to estimate intervention effects and assess whether outcome estimates differ under alternative assessment conditions, while allowing more robust inference through appropriate methods for clustered data and uncertainty estimation.

## Conclusions

This pilot cluster randomized trial found that the adapted six-week IPT-G model was associated with significant reductions in depressive symptoms among women and adolescent girls in Uganda. Although estimated effects were attenuated under bias-minimizing assessment conditions, they remained clinically meaningful. The study also shows that follow-up assessment conditions can materially influence self-reported mental health outcomes, particularly when interventions are delivered through trusted community-based facilitators. Future trials and program evaluations should therefore prioritize independent outcome assessment, clear separation between implementation and research roles and simple procedures that reinforce confidentiality and honest reporting. These safeguards are essential for generating valid evidence on psychological interventions as they move from controlled trials into real-world delivery settings.

## Acknowledgements

We thank the women and adolescent girls who participated in this study and shared their experiences. We also acknowledge the community leaders, Therapy Facilitators, and research assistants who supported participant mobilization, intervention delivery, and follow-up data collection.

We are grateful to Susan Adikini, Rebecca Nabirabwa, Brenda Kalungi, Christine Atugonza, Catherine Akola and Victoria Baranov for their contributions to the implementation and coordination of the study, as well as writing. We further acknowledge the StrongMinds Uganda team for their support throughout study planning and field activities.

## Data Availability Statement

De-identified individual participant data underlying the findings reported in this article, together with the data dictionary and statistical analysis code, will be available after publication to qualified researchers for legitimate scientific purposes. Access is restricted because the dataset contains sensitive mental health and psychosocial information from participants in small rural communities, which may carry a risk of re-identification despite removal of direct identifiers. Requests should be submitted to the Mildmay Research and Ethics Committee, or to the lead author at. Requests will be considered in accordance with the approved ethics protocol, participant consent, and a data-sharing agreement prohibiting re-identification, onward sharing, and non-approved uses of the data. The study protocol will be made available with the article as Supporting Information.

## References

1. GBD 2019 Mental Disorders Collaborators. Global, regional, and national burden of 12 mental disorders in 204 countries and territories, 1990&2013;2019: a systematic analysis for the Global Burden of Disease Study 2019. Lancet Psychiatry. 2022 Feb 1;9(2):137–50. doi:10.1016/S2215-0366(21)00395-3

2. Patel V, Saxena S, Lund C, Thornicroft G, Baingana F, Bolton P, et al. The Lancet Commission on global mental health and sustainable development. The Lancet. 2018 Oct 27;392(10157):1553–98. doi:10.1016/S0140-6736(18)31612-X

3. Kaggwa MM, Najjuka SM, Bongomin F, Mamun MA, Griffiths MD. Prevalence of depression in Uganda: A systematic review and meta-analysis. PLoS One [Internet]. 2022 Oct 20;17(10):e0276552-. Available from: 10.1371/journal.pone.0276552

4. Kaggwa MM, Harms S, Mamun MA. Mental health care in Uganda. Lancet Psychiatry. 2022 Oct 1;9(10):766–7. doi:10.1016/S2215-0366(22)00305-4

5. Akongo D, Gwokyalya V, Tusubira A, Waiswa P, Wamala K, Tuhebwe D, et al. Barriers and facilitators to seeking postpartum depression care services among postpartum mothers in Jinja Hospital Uganda. Journal of Interventional Epidemiology and Public Health. 2024;7(20).

6. Asiimwe R, Nuwagaba-K RD, Dwanyen L, Kasujja R. Sociocultural considerations of mental health care and help-seeking in Uganda. SSM - Mental Health. 2023;4:100232. 10.1016/j.ssmmh.2023.100232

7. World Health Organization. mhGAP intervention guide for mental, neurological and substance use disorders in non-specialized health settings: mental health Gap Action Programme (mhGAP). World Health Organization; 2016.

8. Bolton P, Bass J, Neugebauer R, Verdeli H, Clougherty KF, Wickramaratne P, et al. Group Interpersonal Psychotherapy for Depression in Rural Uganda. JAMA. 2003 Jun 18;289(23):3117. doi:10.1001/jama.289.23.3117

9. Verdeli H, Clougherty K, Bolton P, Speelman L, Lincoln N, Bass J, et al. Adapting group interpersonal psychotherapy for a developing country: experience in rural Uganda. World Psychiatry. 2003;2(2):114.

10. Assefa FB, Tanner L, Kim HY, Platais I, Tindyebwa CM, Kasujja R, et al. Retention and effectiveness of group interpersonal psychotherapy (IPT-G) to treat depression at scale in Uganda and Zambia. BMJ Glob Health. 2025 Nov 16;10(11):e018369. doi:10.1136/bmjgh-2024-018369

11. Kasujja R, Birungi P, Bhamidipati K, Assefa F, Kim HY, Peterson KM, et al. Six-Week Problem Area–Concordant vs 8-Week Problem Area–Discordant Group Interpersonal Psychotherapy. JAMA Netw Open. 2025 Apr 16;8(4):e255242. doi:10.1001/jamanetworkopen.2025.5242

12. Kroenke K, Spitzer RL, Williams JBW. The PHQ-9: validity of a brief depression severity measure. J Gen Intern Med. 2001;16(9):606–13.

13. Rickwood DJ, Coleman-Rose CL. The effect of survey administration mode on youth mental health measures: Social desirability bias and sensitive questions. Heliyon. 2023 Sep;9(9):e20131. doi:10.1016/j.heliyon.2023.e20131

14. Holbrook AL, Green MC, Krosnick JA. Telephone versus Face-to-Face Interviewing of National Probability Samples with Long Questionnaires. Public Opin Q. 2003 Dec;67(1):79–125. doi:10.1086/346010

15. Kempf-Leonard K. Encyclopedia of social measurement. 2004.

16. Tourangeau R, Yan T. Sensitive questions in surveys. Psychol Bull. 2007;133(5):859– 83. doi:10.1037/0033-2909.133.5.859

17. Latkin CA, Edwards C, Davey-Rothwell MA, Tobin KE. The relationship between social desirability bias and self-reports of health, substance use, and social network factors among urban substance users in Baltimore, Maryland. Addictive Behaviors. 2017 Oct;73:133–6. doi:10.1016/j.addbeh.2017.05.005

18. Velloza J, Njoroge J, Ngure K, Thuo N, Kiptinness C, Momanyi R, et al. Cognitive testing of the PHQ-9 for depression screening among pregnant and postpartum women in Kenya. BMC Psychiatry. 2020;20(1):31. doi:10.1186/s12888-020-2435-6

19. Kristófersdóttir KH, Vésteinsdóttir V, Kristjánsdóttir H, Karlsson T, Thorsdottir F. Disclosing depressive symptoms: perceived sensitivity of PHQ-9 items in a general population sample. BMC Psychol. 2026;14(1):294. doi:10.1186/s40359-026-04067-7

20. Uganda Bureau of Statistics. The National Population and Housing Census 2024 – Final Report. Kampala; 2024.

21. Kroenke K, Spitzer RL, Williams JBW, Lowe B. An Ultra-Brief Screening Scale for Anxiety and Depression: The PHQ-4. Psychosomatics. 2009 Nov 1;50(6):613–21. doi:10.1176/appi.psy.50.6.613

22. Löwe B, Unützer J, Callahan CM, Perkins AJ, Kroenke K. Monitoring Depression Treatment Outcomes With the Patient Health Questionnaire-9. Med Care [Internet]. 2004;42(12). Available from: https://journals.lww.com/lww-medicalcare/fulltext/2004/12000/monitoring_depression_treatment_outcomes_with_the.6.aspx

23. Yates CR, M BJ, Arne B, L HA, E SG. Defining Success in Measurement-Based Care for Depression: A Comparison of Common Metrics. 2020 Apr 1;71:312–8. doi:10.1176/appi.ps.201900295

24. Yator O, Mwavua SM, Tele A, Kathono J, Nyongesa V, Amin N, et al. Feasibility of Mini Group Interpersonal psychotherapy (IPT-G mini) for adolescents impacted by COVID-19 in East Africa. medRxiv. 2025 Jan 1;2025.06.13.25329611. doi:10.1101/2025.06.13.25329611

25. Bass J, Neugebauer R, Clougherty KF, Verdeli H, Wickramaratne P, Ndogoni L, et al. Group interpersonal psychotherapy for depression in rural Uganda: 6-month outcomes. British Journal of Psychiatry. 2006 Jun 2;188(6):567–73. doi:10.1192/bjp.188.6.567

26. Jordans MJD, Kohrt BA, Sangraula M, Turner EL, Wang X, Shrestha P, et al. Effectiveness of Group Problem Management Plus, a brief psychological intervention for adults affected by humanitarian disasters in Nepal: A cluster randomized controlled trial. PLoS Med [Internet]. 2021 Jun 17;18(6):e1003621-. Available from: 10.1371/journal.pmed.1003621

27. Musanje K, Atuhumuza E, Kimera E, Mugarura J, Nsereko GM, Mukula HM, et al. How interpersonal psychotherapy groups support recovery from depression: a qualitative study in rural Uganda. Academia Mental Health and Well-Being. 2025 Dec 4;2(4). doi:10.20935/MHealthWellB8022

28. Adong J, Fatch R, Emenyonu NI, Cheng DM, Muyindike WR, Ngabirano C, et al. Social Desirability Bias Impacts Self-Reported Alcohol Use Among Persons With HIV in Uganda. Alcohol Clin Exp Res. 2019 Dec 11;43(12):2591–8. doi:10.1111/acer.14218

29. Espinosa da Silva C, Fatch R, Emenyonu N, Muyindike W, Adong J, Rao SR, et al. Psychometric assessment of the Runyankole-translated Marlowe-Crowne Social Desirability Scale among persons with HIV in Uganda. BMC Public Health. 2024 Jun 19;24(1):1628. doi:10.1186/s12889-024-18886-z

